# Syndromic Surveillance-Based Estimates of Vaccine Efficacy Against COVID-Like Illness from Emerging Omicron and COVID-19 Variants

**DOI:** 10.1101/2021.12.17.21267995

**Authors:** Tanner J. Varrelman, Benjamin Rader, Christina M. Astley, John S. Brownstein

**Affiliations:** Computational Epidemiology Lab, Boston Children’s Hospital, Boston, MA 02115; Department of Epidemiology, Boston University, Boston, MA 02118; Division of Endocrinology, Boston Children’s Hospital, Boston, MA 02115; Harvard Medical School, Boston, MA 02115; Broad Institute of Harvard and MIT, Cambridge, MA 02142

## Abstract

New infections from the omicron variant of SARS-CoV-2 have been increasing dramatically in South Africa since first identification in November 2021. Despite increasing uptake of COVID-19 vaccine, there are concerns vaccine protection against omicron may be reduced compared to other variants. We sought to characterize a surrogate measure of vaccine efficacy in Gauteng, South Africa by leveraging real-time syndromic surveillance data. The University of Maryland Global COVID Trends and Impact Survey (UMD-CTIS) is an online, cross-sectional survey conducted among users sampled from the Facebook active user base. We derived three COVID-like illness (CLI) definitions (stringent, classic, and broad) using combinations of self-reported symptoms (present or not in the prior 24 hours) that broadly tracked with reported COVID-19 cases during June 18, 2021 - December 14, 2021 (inclusive of the delta wave and up-trend of the omicron wave). We used syndromic-surveillance-based CLI prevalence measures among the vaccinated (*P*_*V*_) and unvaccinated (*P*_*U*_) respondents to estimate *V E*_*CLIP*_ = 1 - (*P*_*V*_ /*P*_*U*_), a proxy for vaccine efficacy, during the delta (June 18-July 18, N= 9,387 surveys) and omicron (December 4-14, N= 2,389 surveys) wave periods. We assume no waning immunity, CLI prevalence approximates incident infection with each variant, and vaccinated and unvaccinated survey respondents in the two variant wave periods are exchangeable. The vaccine appears to have consistently lower *V E*_*CLIP*_ against omicron, compared to delta, regardless of the CLI definition used. Stringent CLI (i.e. anosmia plus fever, cough and/or myalgias) yielded a delta *V E*_*CLIP*_ = 0.85 [0.54, 0.95] higher than omicron *V E*_*CLIP*_ = 0.62 [0.46, 0.72]. Classic CLI (cough plus anosmia, fever, and/or myalgias) gave lower estimates (delta *V E*_*CLIP*_ = 0.76 [0.54, 0.87], omicron *V E*_*CLIP*_ = 0.51 [0.42, 0.59]), but omicron was still lower than delta. We acknowledge the potential for measurement, confounding, and selection bias, as well as limitations for generalizability for these self-reported, syndromic surveillance-based *V E*_*CLIP*_ measures. Thus *V E*_*CLIP*_ as estimates of true, population-level vaccine efficacy should therefore be taken with caution. Nevertheless, these preliminary findings demonstrating declining *V E*_*CLIP*_ raise concern for a true decline in vaccine efficacy versus waning immunity as a potential contributor to the omicron variant taking hold in Gauteng and elsewhere.

## 1 Introduction

New infections from the omicron variant of SARS-CoV-2 have been increasing dramatically in South Africa since first identification in November 2021, and despite increasing uptake of COVID-19 vaccine. The majority of confirmed cases have occurred in the Gauteng province [1]. Omicron carries several mutations that potentially contribute to increased transmissibility [2], as well as reduce immune protection. Initial reports show an increased risk of reinfection when facing the omicron variant of the SARS-CoV-2 virus [3]. However, these early claims are based on relatively small sample sizes. Moreover, a recent large scale analysis of COVID-19 test results within South Africa suggests that the current COVID-19 vaccines may not protect as well against the omicron variant [4]. In the setting of novel variants such as omicron, particularly in the early stages of national and international spread, there is a need for preliminary estimates of vaccine efficacy as a contributor to variant emergence and dominance.

To further understand the early emergence of the omicron variant in Gauteng, South Africa, we combine real-time syndromic surveillance data from daily cross-sectional surveys with a COVID-like illness (CLI) prevalence-based vaccine efficacy estimator (*V E*_*CLIP*_).

## 2 Methods

### 2.1 Syndromic Surveillance and Case Data

The University of Maryland Global COVID Trends and Impact Survey (UMD-CTIS), in partnership with Facebook, is a cross-sectional survey that samples Facebook’s active user base, on a daily basis, and collects self-reported information such as demographics, CLI symptoms in the prior 24 hours, and COVID-19 vaccination status (data access and survey questions available https://covidmap.umd.edu) [5,6]. We focused on surveys administered between June 18, 2021 - December 14, 2021 which includes June 18-July 18, 2021 (delta wave period) and December 4 - December 14, 2021 (omicron wave period), from users who self-reported their residing in Gauteng, South Africa (UMD-CTIS data accessed 15 December 2021). These periods were selected to evaluate two temporally separated time periods where there were a sufficient number of confirmed COVID-19 cases (*>* 6,000 cases/day), and sufficient survey data from respondents indicating they were vaccinated with 2-doses (N *>* 200/period). As the UMD-CTIS data is collected and released on a daily basis, in near-real time, the omicron wave period can be extended in real-time as well. Boston Children’s Hospital Institutional Review Board (P00023700) approved this study using UMD-CTIS data. Gautang, South Africa COVID-19 case reports were accessed 15 December, 2021 from the Coronavirus COVID-19 (2019-nCoV) Data Repository and Dashboard for South Africa GitHub [7]. New daily infections were back calculated from the daily cumulative distribution of confirmed cases, and a 7-day rolling average of new daily COVID-19 cases in Gauteng was calculated. Vaccine uptake data for South Africa is available from the Republic of South Africa’s Department of Health [8].

### 2.2 Analytic and Statistical Methods

To understand how breakthrough infections have potentially changed from the delta wave period (“third wave”) to the omicron wave period (“current wave”), we utilize a CLI prevalence based vaccine efficacy estimator (*V E*_*CLIP*_). We developed three CLI definitions to use as a proxy for COVID-19 infection. First we identified symptoms with a notable increase in prevalence in parallel with reported cases during the delta and omicron periods. We then combined these symptoms to create stringent, classic, and broad CLI definitions. We use CLI prevalence (number surveys with self-reported CLI as present in the prior 24 hours, per survey) from the cross-sectional surveys as an estimate of incident symptoms, and CLI symptoms as a proxy for SARS-CoV-2 infection. Finally, we calculate a vaccine efficacy estimate from these syndromic-surveillance data, by comparing the CLI prevalence in the vaccinated and unvaccinated during the delta and omicron wave periods in Gauteng, South Africa (*V E*_*CLIP*_ formula below, confidence interval estimators as per reference [9]):

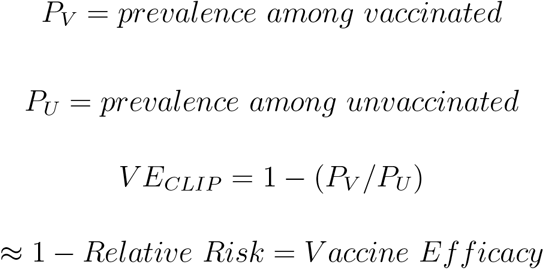

## 3 Results

### 3.1 Derived CLI Definitions

To characterize the constellation of symptoms that can be used to identify COVID-19 within Gauteng, we first identified self-reported symptoms that have appeared to be rising during the delta and omicron waves: cough, fever, and muscle aches and pains (here referred to in short as myalgias). These symptoms were combined with loss of smell or taste (here referred to in short as anosmia), the symptom that is most highly associated with COVID-19 infection throughout the pandemic, and was a consistently strong predictor across countries, time, and syndromic surveillance platforms [10]. These definitions were termed stringent CLI (anosmia with fever, cough, and/or myalgias), classic CLI (cough with anosmia, fever, cough and/or myalgias), and broad CLI (myalgias with anosmia, fever, and/or cough). We illustrate how the prevalence among respondents to UMD-CTIS cross-sectional surveys, using these CLI definitions, track with COVID-19 cases from official case count data, using 7-day moving averages. Figure 1 suggests that the classic CLI definition (cough and at least one of the following: fever, muscle pain, anosmia), provides similar trends to reported cases. This is true across the delta wave time period when the Delta variant was dominant (June to September), and the first two weeks of December when Omicron infections surged. Further, we find that the stringent CLI definition that is designed to be more specific to COVID-19 – but also relies on a less frequently reported symptom (anosmia) – has a weaker signal during the omicron wave period infection than both the classic and broad CLI definitions that include more frequently reported symptoms. Because Omicron cases are rising in Gauteng at present, the signal-to-noise is changing. We therefore include all three CLI definitions for the vaccine efficacy estimates for comparison going forward.

**Figure 1:**
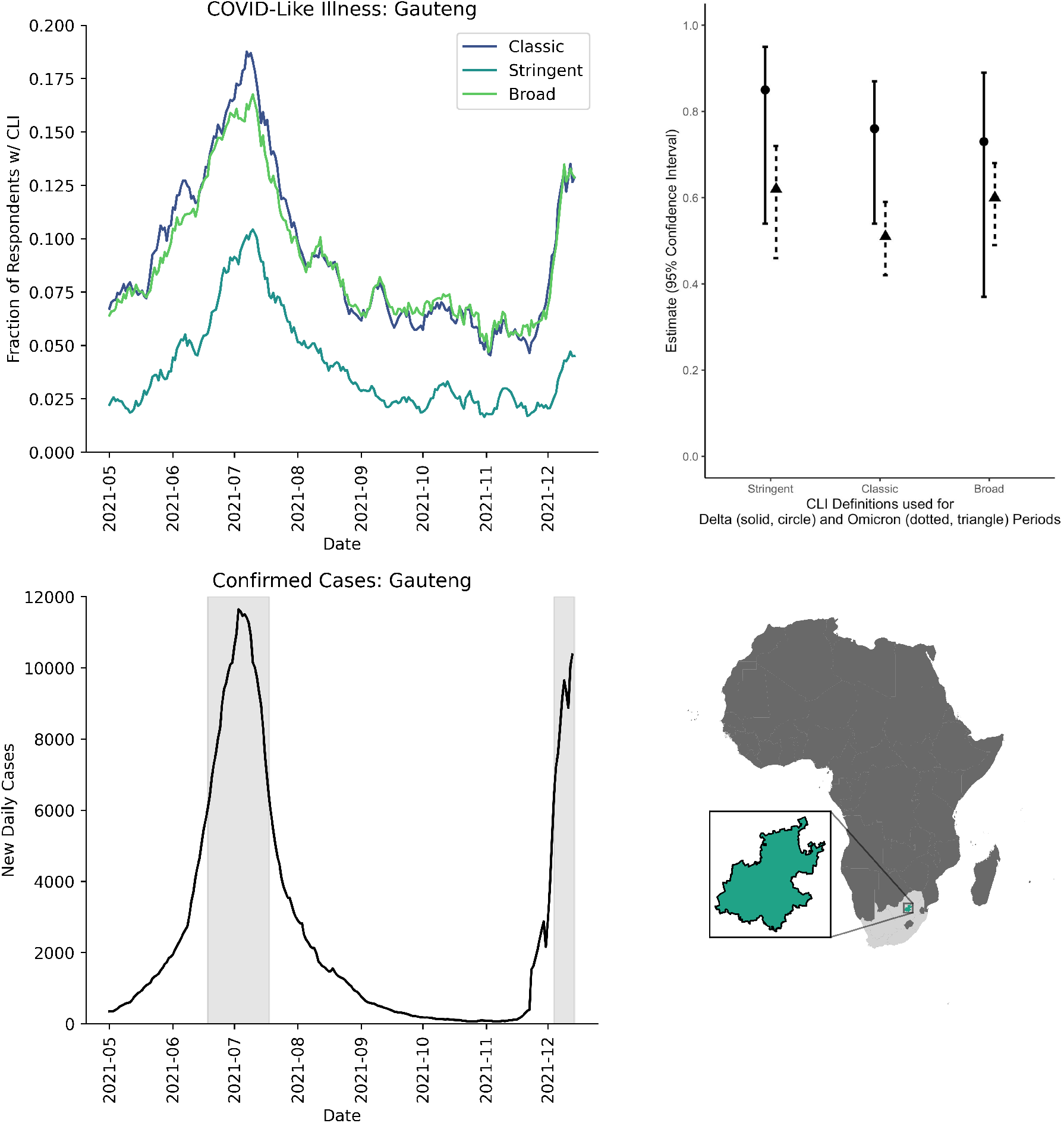
(Top Left) Proportion of UMD-CTIS survey respondents with self-reported COVID-like illness (CLI) in the prior 24 hours using three symptom-based definitions, 1) Stringent CLI: anosmia plus fever, cough, and/or myalgias, 2) Classic CLI: cough plus fever, anosmia, and/or myalgias, and 3) Broad CLI: myalgias plus fever, cough, and/or anosmia, over a 7-day rolling window. (Bottom Left) 7-day rolling average of confirmed new case counts of COVID-19. Gray shaded regions depict the time windows used for delta and omicron comparisons. (Top Right) *V E*_*CLIP*_ estimates and confidence intervals using 3 CLI definitions, and over the two periods. (Top Right) Map showing Africa, South Africa and Gauteng.

### 3.2 CLI to Estimate Vaccine Efficacy

Given the three syndromic surveillance signals based on our CLI definitions tracked with COVID-19 cases, and were thus reasonable proxies for COVID-infection, we then compared CLI among the vaccinated and unvaccinated, during the delta and omicron wave periods. Specifically, we considered vaccinated individuals as those that have self-reported receiving two doses of vaccine, and unvaccinated individuals as those that self-reported having not received any vaccine. Parsing our data this way allows us to leverage classic cohort study theory to approximate the attack rate of COVID-19 for both vaccinated and unvaccinated individuals, using the CLI prevalence among vaccinated and unvaccinated survey respondents to derive *V E*_*CLIP*_ [9]. It is critical to note that our estimate of *V E*_*CLIP*_ is not equivalent to the true vaccine efficacy as we make key assumptions: (1) we assume self-reported prevalent CLI is a valid proxy for incident COVID-19 infection, but breakthrough infections may be asymptomatic and not all CLI is test-positive COVID-19; (2) we assume a simplified model of unvaccinated and vaccinated sub-cohorts, but we do not have information on natural immunity among the unvaccinated, nor on vaccine formulation or timing for the vaccinated; (3) we assume the delta and omicron variants are the sole variants during their respective study periods, and do not account for other variants or viral co-circulation; (4) we assume the survey sampling of the study base (i.e. active Facebook users) is consistent between study periods, non-differential with respect to symptoms, vaccination status and/or potential modifiers of vaccine efficacy. If these assumptions do not hold, the estimator may be upwards or downwards (e.g. omicron infections asymptomatic or cough unrelated to COVID-19, respectively). Therefore these *V E*_*CLIP*_ estimates should be viewed as preliminary measures by which we can compare relative changes in *V E*_*CLIP*_, thereby providing an alert of a potential decline in vaccine efficacy with the emergence of the Omicron variant in Gauteng, South Africa, or elsewhere.

Our syndromic-surveillance informed vaccine efficacy estimates (*V E*_*CLIP*_) show a general pattern of decline from the delta to the omicron wave periods in Gauteng, South Africa (Table 1). Estimates decline within the Gauteng province, regardless of the COVID-like illness definition that we use. Confidence intervals are wide for the delta period, likely due to relative sample size of unvaccinated and vaccinated, as this was early in vaccine roll-out. If Omicron is leading to fewer symptoms per infection, the true efficacy may be even lower – so it is critical to appreciate these syndrome-based definitions would not capture changes in disease severity, hospitalization, or death.

**Table 1:** *V E*_*CLIP*_ calculated for three definitions of CLI (stringent, classic, and broad), across two time periods that capture the spread of the delta and omicron variants.

| **Time Period** | **Delta Dominant Period** | **June 18th, 2021 - July 18th, 2021** | **Omicron Dominant Period** | **Dec. 4th, 2021 - Dec. 13th, 2021** |
| --- | --- | --- | --- | --- |
| **CLI Definition** | **$VE_{CLIP}$** | **$VE_{CLIP}$ 95% CI** | **$VE_{CLIP}$** | **$VE_{CLIP}$ 95% CI** |
| **Stringent CLI:**<br>Anosmia and<br>(Fever or Muscle Pain or Cough) | 0.85 | [0.54, 0.95] | 0.62 | [0.46, 0.72] |
| **Classic CLI:**<br>Cough and<br>(Fever or Muscle Pain or Anosmia) | 0.76 | [0.54, 0.87] | 0.51 | [0.42, 0.59] |
| **Broad CLI:**<br>Muscle Pain and<br>(Fever or Cough or Anosmia) | 0.73 | [0.37, 0.89] | 0.60 | [0.49, 0.68] |

## 4 Discussion

The omicron variant is surging in Gauteng, South Africa, renewing concerns about the ability of current COVID-19 vaccines to protect against novel variants. Utilizing regional data drawn from a global syndromic surveillance platform, we provide early evidence that a relative decline in vaccine efficacy, as estimated by *V E*_*CLIP*_, may be contributing to the omicron variant taking hold in Gauteng, and likely elsewhere. Other investigations ranging from prediction models of genetic changes, analysis of medical databases, and in vitro models have similarly raised concerns about the ability of Omicron to evade vaccine-induced immune response, transmit, and cause disease [2, 4, 11]. Evidence from recent lab based studies suggest that the Omicron variant has the potential to produce more breakthrough cases in 2-dose vaccinated individuals than previous SARS-CoV-2 variants [11], an is consistent with changes that we have observed in our proxy of vaccine efficacy. The South Africa study that analyzed over 200 thousand medical records suggests that the relative vaccine efficacy for omicron is about half that for the delta variant [4], lower than the decline observed using *V E*_*CLIP*_, though the outcome measures in that study are not identical to the symptom-base outcomes utilized here.

This study highlights the value of participatory, remote surveillance as an adjunctive tool for gleaning rapid insights about epidemiologic parameters in the face of changing disease dynamics. Specifically, the UMD-CTIS platform – with its daily study base sampling scheme and broad geographic coverage of thousands of sub-national regions in over a hundred countries worldwide – provides an existing and ongoing data stream for this type of analysis. Previous studies have shown the value of syndromic surveillance platforms for COVID-19 and other infections within specific countries [6, 12]. The use of short surveys delivered to a daily sample of Facebook users globally provides an opportunity to glean initial insights rapidly and on a small geographic scale from an existing data stream, and without the use of regional infrastructure, and with the ability to scale up analyses to other regions as omicron spreads globally.

While these early observational results suggest that UMD-CTIS cross-sectional self-reported symptom and vaccination status reports can be leveraged as a proxy for COVID-19 attack rate in a population with an ongoing epidemic. All three of the CLI definitions support a decline in *V E*_*CLIP*_, and thus a potential decline in vaccine efficacy in the Gauteng province. However, this estimated decline in *V E*_*CLIP*_ can plausibly be attributed to biological mechanisms: 1) increased penetration of the Omicron variant to the COVID-19 vaccine, and/or 2) waning vaccine induced immunity (or confounding by time since vaccination). The present study does not contain enough information to disambiguate between these two alternatives, though UMD-CTIS is updated periodically and may incorporate questions regarding vaccine timing should there be a public health need for these data.

The early results of declining *V E*_*CLIP*_, from the delta wave period to this initial omicron wave period presented here on key assumptions, and should therefore be taken with caution. For instance, our analyses do not account for the vaccine formula that an individual received. This is problematic, as studies have shown varying levels of vaccine efficacy across different vaccine formulations. Further, while our stringent, classic, and broad definitions of CLI can account for potential heterogeneities in COVID-19 symptomatology, the definition is general enough to include seasonal influenza and other febrile respiratory illnesses, as well as non-infectious causes of symptoms. Recent reports of increased influenza throughout the month of November, suggest that influenza and other respiratory pathogens could be contributing to the recent observation of reduced *V E*_*CLIP*_ within Gauteng. Although we have examined individual symptoms and additional definitions of CLI, we find that the use of the classic CLI symptom sets generally captures COVID-19 case trends in Gauteng during this early study period. As cases rise, and the proportion of respondents with anosmia similarly rise, the more specific CLI definition requiring anosmia may be more useful for ensuring COVID-19 specificity, and limiting bias from non-specific symptom trends. With the clinical presentation of Omicron still unknown, the syndromic definitions applied in this study were not used to distinguish between different variants of COVID-19 and instead we referred to outside confirmed reports. Lastly, we are assuming the UMD-CTIS respondents who were vaccinated and unvaccinated during the delta and omicron wave periods can be considered to be exchangeable. Vaccine uptake is not random and the force of infection is heterogeneous; those with higher risk and higher means may get vaccine earlier and have immunity that wanes earlier, while vaccinated individuals may be both healthier or more willing to engage in social or unmasked activities. UMD-CTIS is a statistical sample of Facebook users, and there may be important limitations to generalizing effect estimates from a selected sample [13]. Further studies could address this by incorporating covariates – such as gender, age, illness duration, and the potential risk behaviors of survey respondents – to fully understand the potential biases, and better characterize the potential risks of emerging virus variants.

While the syndromic surveillance informed proxy of vaccine efficacy may not be the gold standard for estimating vaccine efficacy, we have shown that analysis of regional data from UMD-CTIS can quickly point towards high-level changes leveraging remote, real-time CLI trends survey respondents across time. Still, this initial report has only described a very limited set of questions that can be asked using UMD-CTIS to better understand the epidemiological profile of Omicron in Gauteng, South Africa and globally. As we continue to face SARS-CoV-2 variants, and seasonal and emerging infectious disease in general, it is crucial that we combine insights from lab-based studies, with early insights from syndromic surveillance data streams.

## Data Availability

Access to the CTIS data can be can be requested from Facebook Data for Good website (https://dataforgood.facebook.com/dfg/docs/covid-19-trends-and-impact-survey-request-for-data-access). The UMD Global CTIS Open Data API, Microdata Repository, and contingency tables are available from The University of Maryland Social Data Science Center Global COVID-19 Trends and Impact Survey website (https://covidmap.umd.edu).

## Acknowledgement

This work was supported by a Facebook Sponsored Research Agreement (TJV, BR, CMA, JSB, INB1116217). The funding sources played no role in the study design, collection, analysis, interpretation, writing or decision to submit the paper for publication.

## References

[1] Ewen Callaway and Heidi Ledford. How bad is omicron? what scientists know so far. Nature News, 2021.

[2] Bingrui Li, Xin Lu, Kathleen M. McAndrews, and Raghu Kalluri. Mutations in the spike rbd of sars-cov-2 omicron variant may increase infectivity without dramatically altering the efficacy of current multi-dosage vaccinations. bioRxiv, 2021.

[3] Juliet R.C. Pulliam, Cari van Schalkwyk, Nevashan Govender, Anne von Gottberg, Cheryl Cohen, Michelle J. Groome, Jonathan Dushoff, Koleka Mlisana, and Harry Moultrie. Increased risk of sars-cov-2 reinfection associated with emergence of the omicron variant in south africa. medRxiv, 2021.

[4] Discovery health, south africa’s largest private health insurance administrator, releases at-scale, real-world analysis of omicron outbreak based on 211,000 covid-19 test results in south africa, including collaboration with the south africa [press release]. Dec 2021.

[5] Frauke Kreuter. Partnering with a global platform to inform research and public policy making. Survey Research Methods, 14(2):159–163, jun 2020.

[6] Christina M. Astley, Gaurav Tuli, Kimberly A. Mc Cord, Emily L. Cohn, Benjamin Rader, Tanner J. Varrelman, Samantha L. Chiu, Xiaoyi Deng, Kathleen Stewart, Tamer H. Farag, Kristina M. Barkume, Sarah LaRocca, Katherine A. Morris, Frauke Kreuter, and John S. Brownstein. Global monitoring of the impact of the covid-19 pandemic through online surveys sampled from the facebook user base. Proceedings of the National Academy of Sciences, 118(51), 2021.

[7] Vukosi Marivate and Herkulaas MvE Combrink. Use of available data to inform the covid-19 outbreak in south africa: A case study. Data Science Journal, 19(1):1–7, 2020.

[8] Covid-19 online resource & news portal sacoronavirus.co.za, 2021.

[9] AW Hightower, WA Orenstein, and SM Martin. Recommendations for the use of taylor series confidence intervals for estimates of vaccine efficacy. Bull World Health Organ, 66(1):99–105, 1988.

[10] Carole H Sudre, Ayya Keshet, Mark S Graham, Amit D Joshi, Smadar Shilo, Hagai Rossman, Benjamin Murray, Erika Molteni, Kerstin Klaser, Liane D Canas, Michela Antonelli, Long H Nguyen, David A Drew, Marc Modat, Joan Capdevila Pujol, Sajaysurya Ganesh, Jonathan Wolf, Tomer Meir, Andrew T Chan, Claire J Steves, Tim D Spector, John S Brownstein, Eran Segal, Sebastien Ourselin, and Christina M Astley. Anosmia, ageusia, and other COVID-19-like symptoms in association with a positive SARS-CoV-2 test, across six national digital surveillance platforms: an observational study. The Lancet Digital Health, 3(9):e577.#x2013;e586, sep 2021.

[11] Wanwisa Dejnirattisai, Robert H Shaw, Piyada Supasa, Chang Liu, Arabella S V Stuart, Andrew J Pollard, Xinxue Liu, Teresa Lambe, Derrick Crook, Dave I Stuart, Juthathip Mongkolsapaya, Jonathan S Nguyen-Van-Tam, Matthew D Snape, Gavin R Screaton, and Com-COV2 Study Group. Reduced neutralisation of sars-cov-2 omicron-b.1.1.529 variant by post-immunisation serum. medRxiv, 2021.

[12] Arjuna Maharaj, Jennifer Parker, Jessica Hopkins, Effie Gournis, I. Bogoch, Benjamin Rader, Christina Astley, Noah Ivers, Jared Hawkins, Nancy Vanstone, Ashleigh Tuite, David Fisman, John Brownstein, and Lauren Lapointe-Shaw. The effect of seasonal respiratory virus transmission on syndromic surveillance for covid-19 in ontario, canada. The Lancet Infectious Diseases, 21, 03 2021.

[13] Valerie C. Bradley, Shiro Kuriwaki, Michael Isakov, Dino Sejdinovic, Xiao-Li Meng, and Seth Flaxman. Unrepresentative big surveys significantly overestimated us vaccine uptake. Nature, 2021.

